# Risk Prediction Modelling of 30-day all-cause mortality following percutaneous coronary intervention in an Australian population: Leveraging Machine Learning

**DOI:** 10.1101/2025.03.01.25323134

**Authors:** Mohammad Rocky Khan Chowdhury, Diem Dinh, Angela Brennan, Christopher M. Reid, Shane Nanayakkara, Jeffrey Lefkovits, Derek P. Chew, Md Nazmul Karim, Mohammad Ali Moni, Md Shofiqul Islam, Baki Billah, Dion Stub

## Abstract

**Background:** Pre-procedural risk prediction of 30-day all-cause mortality after percutaneous coronary intervention (PCI) aids in clinical decision-making and benchmarking hospital performance. This study aimed to identify pre-procedural factors to predict the risk of 30-day all-cause mortality post-PCI using machine learning (ML) approaches.

**Methods:** The study analysed 93,055 consecutive PCI procedures. Boruta feature selection method was used to identify key predictive variables. Seven ML algorithms were employed for models’ development and validation. Model performance was assessed using standard metrics for validation dataset. SHapley Additive exPlanations (SHAP) method was used to explain leading predictive variables.

**Results:** Among the seven ML algorithms, the Extreme Gradient Booster (XGB) had the better performance across most metrics, such as accuracy (86.7%), root mean square error (36.5%), specificity (82.5%), precision (54.0%), F1 score (52.7%), and Brier score (13.3%). The XGB model also demonstrated strong discriminatory power, achieving a receiver operating characteristics-area under the curve (ROC-AUC) of 85.5% (95% CI: 83.5%–87.4%). The XGB model identified left ventricular ejection fraction (LVEF), acute coronary syndrome (ACS), estimated glomerular filtration rate (eGFR), age, and complex lesion as the five leading factors associated with 30-day mortality post-PCI. Other factors, in order, were cardiogenic shock, body mass index (BMI), intubated out-of-hospital cardiac arrest (OHCA), lesion location, mechanical ventricular support, gender, and peripheral vascular disease (PVD).

**Conclusion:** The XGB algorithm was identified as the best predictive model for 30-day all-cause mortality post-PCI. It is essential to underscore the need for further validation of the model with external data to ensure its applicability to other populations.

**WHAT IS ALREADY KNOWN ON THIS TOPIC:**

- risk-adjustment model for an Australian percutaneous coronary intervention (PCI) patient population was previously developed to predict 30-day mortality post-PCI using traditional regression model.
- knowledge, patient characteristics, and clinical practices evolve over time, requiring frequent model updates to reflect new evidence, guidelines, and interventions

**WHAT THIS STUDY ADDS:**

- A machine learning (ML)-based preprocedural risk prediction model for 30-day mortality post-PCI was developed. The Extreme Gradient Booster (XGB) model was identified as the top performer in predicting 30-day all-cause mortality post-PCI. The model selected left ventricular ejection fraction, acute coronary syndrome, estimated glomerular filtration rate, age, and complex lesion as the top influential factors.

**HOW THIS STUDY MIGHT AFFECT RESEARCH, PRACTICE OR POLICY:**

- Risk prediction models aid clinical decision-making, enhance patient counselling, improve care quality, inform healthcare policies, and advance research.

## INTRODUCTION

Percutaneous coronary intervention (PCI) is one of the most widely performed medical procedures,^1^ and is a highly effective in treating coronary artery disease ^2^. Whilst PCI is extremely safe, patients remain at risk of mortality.^3^ In 2020-2021, approximately 48,000 PCIs were performed in Australia, with 75% being performed in males.^4^ It is worth noting that similar to other cardiac procedures, approximately two-thirds of deaths after PCI occur within the first 30 days following the procedure.^5^ In Australia, the prevalence of 30-day all-cause mortality post-PCI was around 2%.^6^ The 30-day mortality is influenced by various factors including patient demographic and their pre-procedural clinical status.^7–10^ Therefore, in order to assess pre-procedural risk of 30-day all-cause mortality, it is essential to identify and understand it’s related factors.

A risk-adjusted model for predicting 30-day all-cause mortality post-PCI can aid physicians in selecting optimal interventions tailored to individual demographics, lifestyles, patient comorbidities, and clinical presentations. Such models are also essential when comparing outcomes across individual institutions and regions. Thus, this approach may enhance intervention quality by improving risk adjustment and subsequently may reduce post-PCI mortality as well as costs through cost-effective care strategies.^11^ ^12^ While existing risk-adjusted models developed by clinical quality registries have been valuable, their applicability to contemporary PCI populations may be limited due to variations in study populations, healthcare systems, registry uniformity, risk factors, and methodological approaches.^7–10^ Additionally, many models in the literature traditionally rely on multivariable logistic and Cox’s regression methods for outcome prediction.^7–10^ ^13^

In recent years, there has been a growing interest in the use of Machine Learning (ML) methods in developing risk prediction models.^14–16^ Traditional methods (e.g. logistic regression or Cox regression) require more structural data, greater human input for the verification of distributional assumptions and incorporation of application knowledge in choosing the input parameters.^17^ Conversely, ML approaches deal with high-level non-structural big data from patient databases. These are often able to detect sophisticated data patterns with a multitude of variables that can be tested for numerous interactions and nonlinear relationships with the outcome that traditional statistical methods are sometimes struggle to explain.^18–20^ Further, these approaches have been successfully applied to predict patient prognoses in many public health areas, such as the risk of readmission after hospital discharge, cancer progression, and diabetic complications.^21–23^ Current evidence indicated that ML methods outperformed traditional regression models in population-specific mortality studies.^24–26^ Though the application of ML has amplified in medical and health care in Australia, its potential applications in 30-day all-cause mortality post-PCI has not been extensively explored in a contemporary Australian population. Therefore, this current study aims to identify pre-procedural factors associated with 30-day all-cause mortality post-PCI for an Australian population, find the best ML approach and compare ML’s performance metrics with the traditional logistic regression method.

## METHODS

### Study Population

Data used in this study were collected by the Victorian Cardiac Outcomes Registry (VCOR) comprising 93,055 consecutive PCI cases from 33 (15 public and 18 private) participating Victorian (a state in Australia) hospitals between 1 January 2013 to 31 December 2021. Patient-centred demographics, comorbidities, procedural details, in-hospital and 30-day mortality were captured by the registry, where each PCI was treated as a separate observation. PCI procedures were excluded if they were not the index admission or had missing outcome measures.^27^

### Outcome variable

The outcome variable for this study was 30-day all-cause mortality post-PCI.

### Selection of potential factors of 30-day morality

An recent systematic review identified 17 key variables as significantly associated with 30-day mortality post-PCI, however only 11 of them were available in the VCOR.^28^ The available variables are: age, gender, PCI indication or acute coronary syndrome (ACS), cardiogenic shock, intubated out-of-hospital cardiac arrest (OCHA), left ventricular ejection fraction (LVEF), estimated glomerular filtration rate (eGFR), mechanical ventilation, history of diabetes, vascular diseases (peripheral vascular disease (PVD), and cerebrovascular disease (CVD)). The following five variables however were not available in the VCOR registry: hypertension, single or multivessel diseases, heart failure, Thrombolysis in Myocardial Infarction (TIMI) flow and urgency of PCI. The percutaneous entry location is determined by the physician’s decision, making it inappropriate for risk prediction, and was excluded from modelling. The following variables: body mass index (BMI), chronic total occlusion (CTO), lesion complexity (ACC/AHA Lesion Classification B2/C), previous coronary artery bypass grafting (CABG) and lesion location were not among the proposed 17 variables, however they were included in the list of potential factors based on consultation with expert interventional cardiologists. Operational definitions for all of these variables were presented in Supplementary Table S1.

### Handling of missing data

A higher proportion of missing values was observed for LVEF (11.3%), followed by eGFR (7.5%) (Table 1). The missing values for individual variables are all below 15%, indicating the feasibility of performing multiple imputations.^29^ Missing values in this study were imputed using Multiple Imputations by Chained Equations (MICE) with fully conditional specification.^30^

**Table 1.** Background profile and 30-day mortality rate post-PCI.

| Factors | Number (%) | 30-day mortality |  |  |
| --- | --- | --- | --- | --- |
|  |  | Survival | Mortality | p-value |
| **Age group** |  |  |  |  |
| <80 years | 79,629 (85.6) | 78,270 (98.3) | 1,359 (1.7) | <0.001 |
| ≥80 years | 13,426 (14.4) | 12,846 (95.7) | 480 (4.3) |  |
| **Gender** |  |  |  |  |
| Male | 70,713 (76.0) | 69,344 (98.1) | 1,369 (1.9) | <0.001 |
| Female | 22,342 (24.0) | 21,772 (97.4) | 570 (2.6) |  |
| **Body mass index** |  |  |  |  |
| Underweight | 649 (0.7) | 649 (95.8) | 30 (4.6) | <0.001 |
| Normal | 21,250 (22.8) | 20,665 (97.3) | 585 (2.7) |  |
| Overweight | 37,538 (40.3) | 36,854 (98.2) | 684 (1.8) |  |
| Obese | 32,639 (35.1) | 32,138 (98.5) | 501 (1.5) |  |
| Missing | 979 (1.1) |  |  |  |
| **PCI indication** |  |  |  |  |
| ST-elevated myocardial infraction | 19,069 (20.5) | 17,776 (93.2) | 1,293 (6.8) | <0.001 |
| Non-ST-elevated myocardial infraction | 21,795 (23.4) | 21,475 (98.5) | 320 (1.5) |  |
| Unstable angina | 6,106 (6.6) | 6,076 (99.5) | 30 (0.5) |  |
| Non-ACS | 46,085 (49.5) | 45,789 (99.4) | 296 (0.6) |  |
| **Cardiogenic shock** |  |  |  |  |
| No | 90,960 (97.8) | 89,950 (98.9) | 1,010 (1.1) | <0.001 |
| Yes | 2,094 (2.2) | 1,165 (55.6) | 929 (44.4) |  |
| Missing | 1 (0.0) |  |  |  |
| **Intubated OHCA** |  |  |  |  |
| No | 91,999 (98.9) | 90,562 (98.4) | 1,437 (1.6) | <0.001 |
| Yes | 1056 (1.1) | 554 (52.5) | 502 (47.5) |  |
| **LVEF** |  |  |  |  |
| Normal | 56,053 (60.3) | 55,736 (99.4) | 317 (0.6) | <0.001 |
| Mild | 14140 (15.2) | 13,892 (98.3) | 248 (1.7) |  |
| Moderate | 8238 (8.8) | 7,839 (95.2) | 399 (4.8) |  |
| Severe | 4070 (4.4) | 3,452 (84.2) | 618 (15.2) |  |
| Missing | 10,554 (11.3) |  |  |  |
| **eGFR** |  |  |  |  |
| Normal | 38,737 (41.6) | 38,456 (99.3) | 281 (0.7) | <0.001 |
| Mild | 28,795 (30.9) | 28,327 (98.4) | 468 (1.6) |  |
| Moderate | 16,146 (17.4) | 15,441 (95.6) | 704 (4.4) |  |
| Severe | 2440 (2.6) | 2,183 (89.5) | 257 (10.5) |  |
| Missing | 6,937 (7.5) |  |  |  |
| **Mechanical ventilation** |  |  |  |  |
| No | 92,348 (99.2) | 90,727 (98.2) | 1,621 (1.8) | <0.001 |
| Yes | 705 (0.8) | 387 (54.9) | 318 (45.1) |  |
| Missing | 2(0.0) |  |  |  |
| **Diabetes** |  |  |  |  |
| No | 71,928 (77.3) | 70,511 (98.0) | 1,417 (2.0) | <0.001 |
| Yes | 21,122 (22.7) | 20,603 (97.5) | 519 (2.5) |  |
| Missing | 5 (0.01) |  |  |  |
| **Peripheral vascular disease** |  |  |  |  |
| No | 89,806 (96.5) | 88,016 (98.0) | 1,790 (2.0) | <0.001 |
| Yes | 3242 (3.5) | 3,098 (95.6) | 144 (4.4) |  |
| Missing | 7 (0.01) |  |  |  |
| **Cerebrovascular disease** |  |  |  |  |
| No | 89,729 (96.4) | 87,940 (98.1) | 1,789 (2.0) | <0.001 |
| Yes | 3319 (3.6) | 3,173 (95.6) | 146 (4.4) |  |
| Missing | 7 (0.01) |  |  |  |
| **Chronic total occlusion** |  |  |  |  |
| No | 89,473 (96.1) | 87,646 (98.0) | 1,827 (2.0) | <0.001 |
| Yes | 3582 (3.9) | 3,470 (96.9) | 112 (3.1) |  |
| **Previous PCI** |  |  |  |  |
| No | 62,317 (67.0) | 60,793 (97.5) | 1,524 (2.5) | <0.001 |
| Yes | 30,734 (33.0) | 30,321 (98.7) | 413 (1.3) |  |
| Missing | 4 (0.0) |  |  |  |
| **Previous CABG** |  |  |  |  |
| No | 86,437 (92.9) | 84,632 (97.9) | 1,805 (2.1) | 0.642 |
| Yes | 6614 (7.1) | 6,482 (98.0) | 132 (2.0) |  |
| **Complex lesion** |  |  |  |  |
| Lesion A or B1 | 37,360 (40.1) | 36,901 (98.8) | 459 (1.2) | <0.001 |
| Lesion B2 or C | 55,659 (59.9) | 54,215 (97.3) | 1480 (2.7) |  |
| **Lesion location** |  |  |  |  |
| Right coronary artery | 29,436 (31.6) | 28,883 (98.1) | 553 (1.9) | <0.001 |
| Left anterior descending | 39,982 (43.0) | 39,129 (97.9) | 853 (2.1) |  |
| Circumflex artery | 20,509 (22.0) | 20,177 (98.4) | 332 (1.6) |  |
| Left main | 1,618 (1.7) | 1,457 (90.0) | 161 (8.0) |  |
| Graft | 1,510 (1.6) | 1,470 (97.3) | 40 (2.7) |  |
| **Total** | 93,055 (100.0) | 91,116 (97.9) | 1,939 (2.1) |  |

Note: CABG, coronary artery bypass grafting; eGFR, estimated glomerular filtration rate; LVEF, Left Ventricular Ejection Fraction; OHCA, out-of-hospital cardiac arrest; PCI, percutaneous coronary intervention

### Analyses

Patients’ characteristics were reported using mean+/-standard deviation (SD) or median and percentiles for numerical data, where apply, and percentages for categorical data. The multicollinearity was assessed by the Variance Inflation Factor (VIF) (VIF>10 indicates multicollinearity) (Supplementary Figure S1).^31^ Further, first-degree interaction effect between clinically relevant variables were also investigated. A Chi-square test was performed to assess the association between 30-day all-cause mortality status and the independent variables. A p-value ≤0.05 or less was considered as statistical significance.

The Boruta feature selection method was used to make a short list of the plausible variables of 30-day all-cause mortality. Boruta, a wrapper-based method using the random forest classifier, is known for its consistency and lack of bias, making it superior to other variable selection techniques.^32^ Seven ML models were developed using the short-listed variables. The most influential variables were identified based on the best-performing models and explained using the SHapley Additive exPlanations (SHAP) method.^33^ All data analyses were undertaken using Stata (version 18/ StataCorp LLC), and Python (version 3.12.2) statistical software packages.

### ML model development and selection of best ML model

Based on previous literature,^34–36^ seven ML algorithms were employed to develop predictive models, which included Decision Tree (DT), Extreme Gradient Boosting (XGB), Gradient Boosting (GB), Linear Discriminant Analysis (LDA), Logistic Regression (LR), Random Forest (RF), and Stochastic Gradient Boosting (SGB) (Supplementary Table S2).

The data was split by 70% and 30%. The models were developed in 70% of entire dataset (training dataset) and rest of 30% data was used for validation (test dataset). The class imbalance of outcome was addressed using Adaptive Synthetic (ADASYN) resampling technique. Each training model was optimized with hyperparameter tuning using 10-fold cross validation protocol (Supplementary Table S3). Finally, the training models were validated in 30% validation dataset. The best ML model was selected based on the comparison of performance metrics (accuracy, root mean square error (RMSE), sensitivity/recall, specificity, precision, F1 score, receiver operating characteristics – area under the curve (ROC-AUC) curve with 95% confidence interval (CI), precision-recall (PR) curve with 95% CI, Brier score and calibration plot) in validation dataset (Supplementary Table S4). Schematic presentation of best ML model selection was shown in Figure 1.

**Figure 1.**
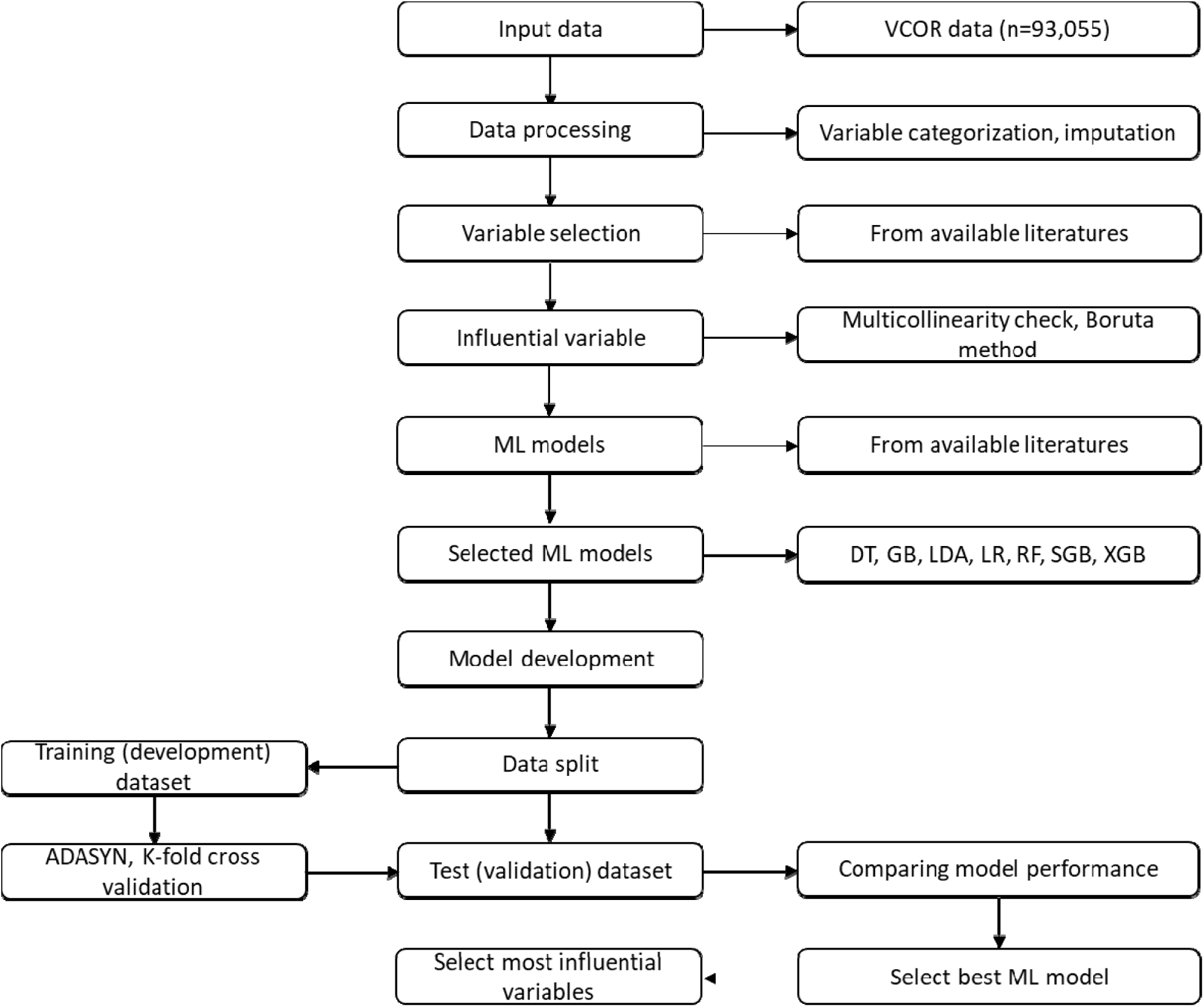
Schematic presentation of machine learning model development Note: VCOR, Victorian Cardiac Outcome Registry; ML, Machine Learning; DT, Decision Tree; GB, Gradient Boosting; LDA, Linear Discriminant Analysis; LR, Logistic Regression; RF, Random Forest; SGB, Stochastic Gradient Boosting; XGB, Extreme Gradient Boosting; ADASYN, Adaptive Synthetic resampling.

### Health services performance

The performance of individual hospitals regarding patients’ adjusted risk of 30-day all-cause mortality is depicted in funnel plots. Funnel plots were utilized as a visual tool for comparing health services by plotting estimates of risk-adjusted 30-day all-cause mortality rates against the total number of procedures performed.^37^ These plots are a valuable addition to performance monitoring systems. To generate the funnel plot, adjusted risk was computed for each patient using regression coefficients derived from traditional LR analysis with the variables identified by the best ML method.

## RESULTS

### Baseline patient characteristics

An overview of the baseline characteristics of the 93,055 patients is presented in Table 1. The average age was 66.5 (±11.9) years and 76% were male. Around 50.5% of patients presented with ACS and 22.7% had a prior history of diabetes. CVD was present in 3.6% and PVD was present in 3.5% patients. The rate of intubated OHCA was 1.1%, and 2.2% presented with cardiogenic shock, and 2.6% of patients had severe renal impairment (eGFR <30 mL/min/1.73 m^2^).

The overall 30-day all-cause mortality was 2.1%. Patients who presenting with cardiac arrest had the highest mortality rates (47.5%) followed by cardiogenic shock (44.1%). Further, patients with severely reduced ejection fraction (LVEF<30%) had a mortality rate of 15.2%, while those with severe renal impairment (eGFR<30 mL/min/1.73 m^2^) had a mortality of 10.5%. The mortality in elderly patients (80 years and over) was 4.3% while those with ST-elevated myocardial infraction (STEMI) had a mortality rate of 6.8% (Table 1).

### Potential influential factors

The Boruta feature selection method identified following eight key factors influencing 30-day all-cause mortality post-PCI: LVEF, cardiogenic shock, ACS, intubated OHCA, eGFR, mechanical ventricular support, age and complex lesion. The factors BMI, PVD, lesion location and gender were selected as moderate influential. The factors CVD, CTO, diabetes, previous PCI and previous CABG were identified as having minimal or no impact on the mortality (Figure 2).

**Figure 2.**
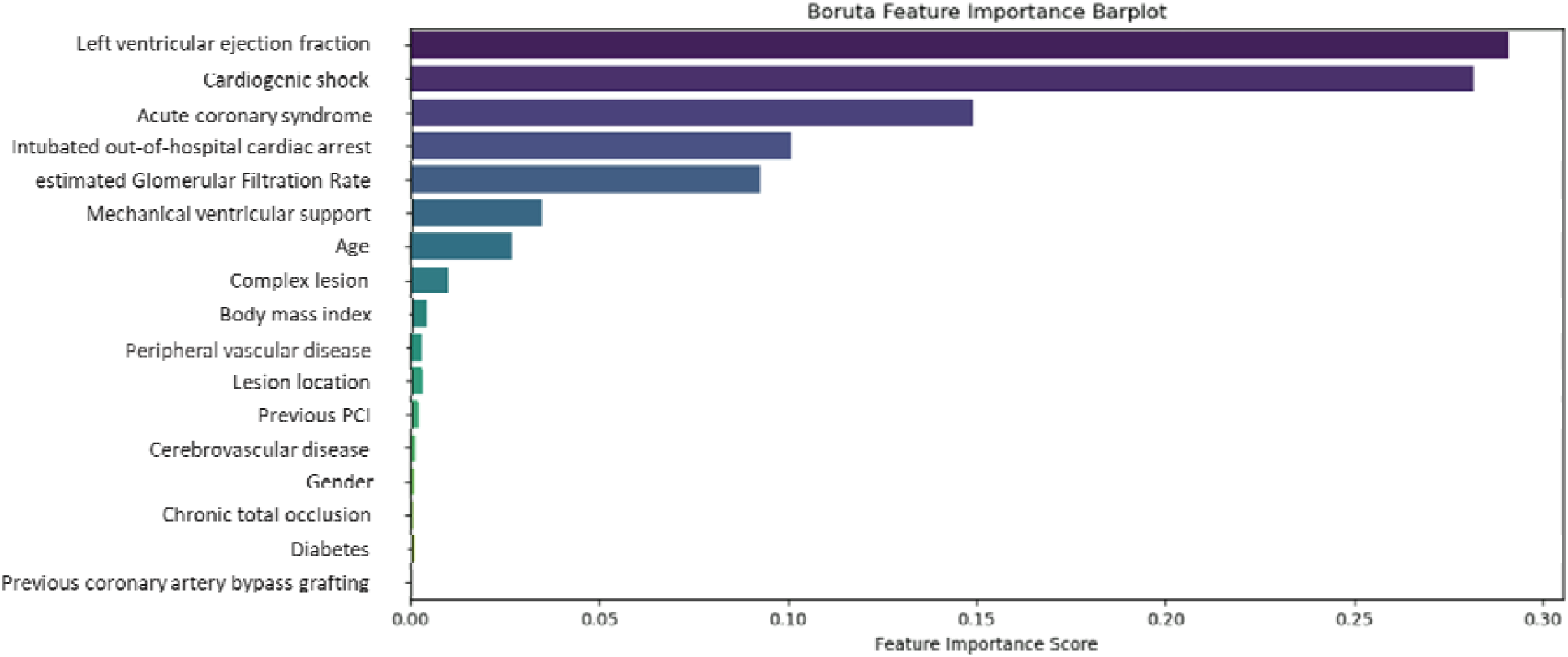
Boruta feature selection method to identify influential factors.

### ML model selection

Twelve influential factors, including sex, which was forcibly included due to its significance,^38^ were used to assess the prediction performance of each of the ML models. Among the seven ML models, the XGB model demonstrated slightly better performance across most metrics, including accuracy (86.7%), RMSE (36.5%), specificity (82.5%), precision (54.0%), F1 score (52.7%), and Brier score (13.3%) (Table 2). The XGB model demonstrated satisfactory performance across other metrics as well, which include a sensitivity/recall of 76.5%, a ROC-AUC of 85.5% (95% CI: 83.5%–87.4%), a PR score of 31.4% (95% CI: 27.1%–35.5%), and improved calibration (Table 2 and Figure 3).

**Figure 3.**
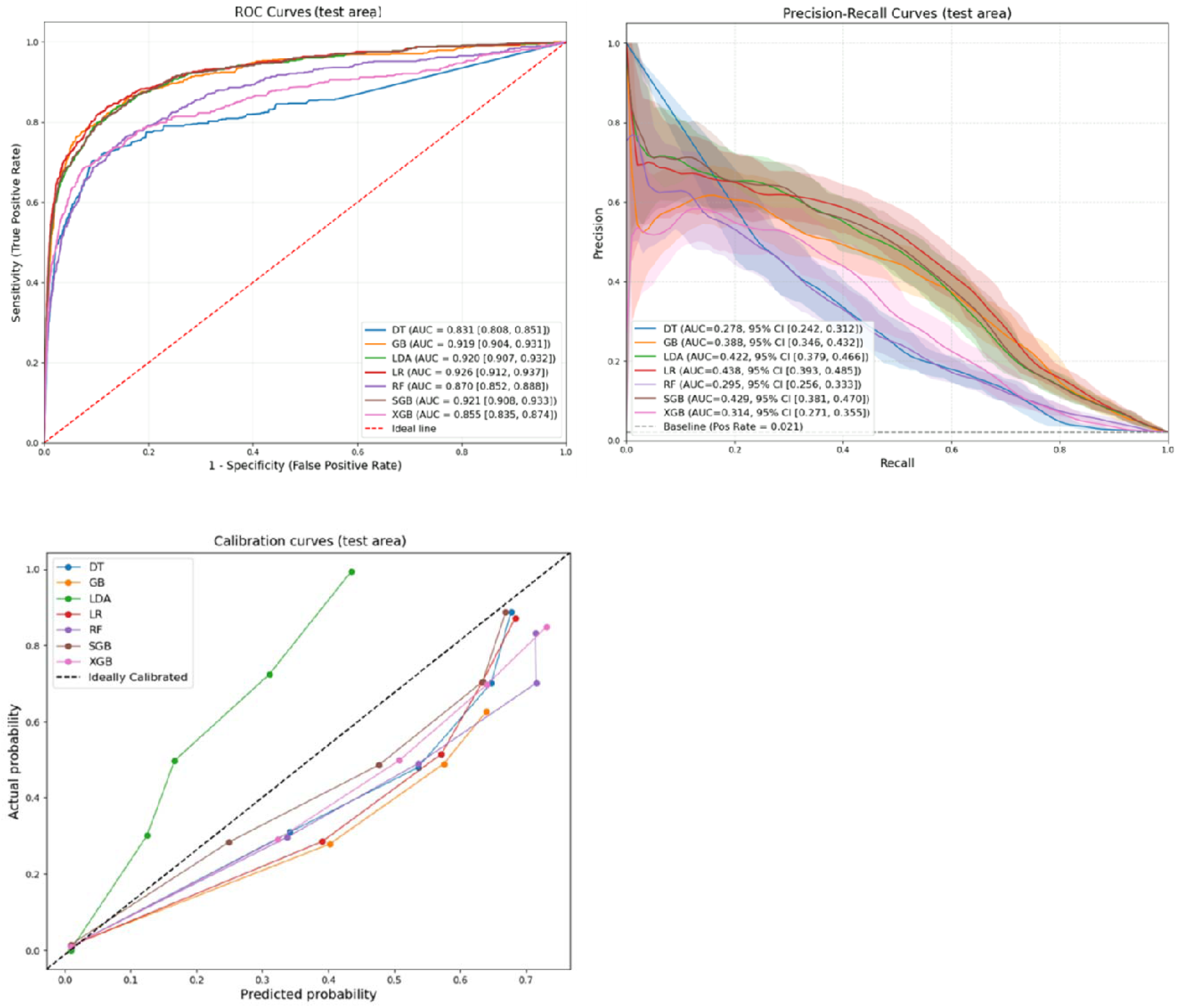
Receiver operating characteristics-area under the curve (ROC-AUC) curve, precision-recall curve and calibration curve for ML models Note: CI, confidence interval; DT, Decision Tree; GB, Gradient Boosting; LDA, Linear Discriminant Analysis; LR, Logistic Regression; RF, Random Forest; SGB, Stochastic Gradient Boosting; XGB, Extreme Gradient Boosting. For both ROC and PR scores, higher values indicate better model performance

**Table 2.** Models’ performance (test/validation dataset) in predicting 30-day mortality post-PCI.

| Models | Performance metrics of models |  |  |  |  |  |  |
| --- | --- | --- | --- | --- | --- | --- | --- |
|  | Accuracy | RMSE | Sensitivity/recall | Specificity | Precision | F1 score | Brier score |
| **DT** | 0.856 | 0.380 | 0.756 | 0.815 | 0.537 | 0.524 | 0.144 |
| **GB** | 0.842 | 0.398 | 0.893 | 0.761 | 0.533 | 0.450 | 0.158 |
| **LDA** | 0.835 | 0.405 | 0.895 | 0.767 | 0.536 | 0.503 | 0.165 |
| **LR** | 0.830 | 0.413 | 0.912 | 0.748 | 0.535 | 0.494 | 0.171 |
| **RF** | 0.853 | 0.384 | 0.792 | 0.801 | 0.536 | 0.515 | 0.147 |
| **SGB** | 0.824 | 0.419 | 0.921 | 0.733 | 0.533 | 0.486 | 0.175 |
| **XGB** | 0.867 | 0.365 | 0.765 | 0.825 | 0.540 | 0.527 | 0.133 |

Note: RMSE, root mean square error; DT, Decision Tree; XGB, Extreme Gradient Boosting; GB, Gradient Boosting; LDA, Linear Discriminant Analysis; LR, Logistic Regression; RF, Random Forest; SGB, Stochastic Gradient Boosting
For Accuracy, Sensitivity, Specificity, Precision and F1 score, higher values indicate better model performance. For RMSE and Brier score lower values indicate better model performance

For sensitivity/recall, the SGB model outperformed the others (92.1%) (Table 2). In terms of discrimination power, the ML based LR model achieved the highest ROC-AUC score of 92.6% (95% CI: 91.2% - 93.7%) compared to 85.5% (95% CI: 83.5%–87.4%) of XGB model. The calibration curve for SGB model showed better calibration compared to other models (Figure 3). Also, ML based LR model achieved the highest PR score (43.8%, 95% CI: 39.3 – 48.5) (Figure 3). Furthermore, Supplementary Table S5 and Supplementary Figure S2 showed models’ performance for the training data set.

### Sensitivity analysis

In the sensitivity analysis, all models were redeveloped and their ROC-AUC scores were evaluated for validation dataset without addressing class imbalance. Additionally, ROC-AUC was assessed using a dataset with missing values removed and without addressing class imbalance. In both cases, XGB model outperformed other ML models in prediction 30-day all-cause mortality post PCI. While ROC-AUC was assessed using a dataset with missing values removed and addressing class imbalance, LR model outperformed other ML models. Also, the XGB model demonstrated satisfactory performance with ROC-AUC score of 76.6% (95% CI: 74.0% - 79.6%) (Supplementary Figure S3).

### Interpretability of top factors in SHAP plot of the XBG model

The SHAP plot in the Figure 4 provides insights into the influence and direction of various variables on predicting 30-day all-cause mortality post-PCI, as depicted by the distribution of red and blue dots. The five leading factors identified by the XGB model were LVEF, ACS, eGFR, age, and complex lesion. Other factors, in order, were cardiogenic shock, BMI, intubated OHCA, lesion location, mechanical ventricular support, gender, and PVD.

**Figure 4.**
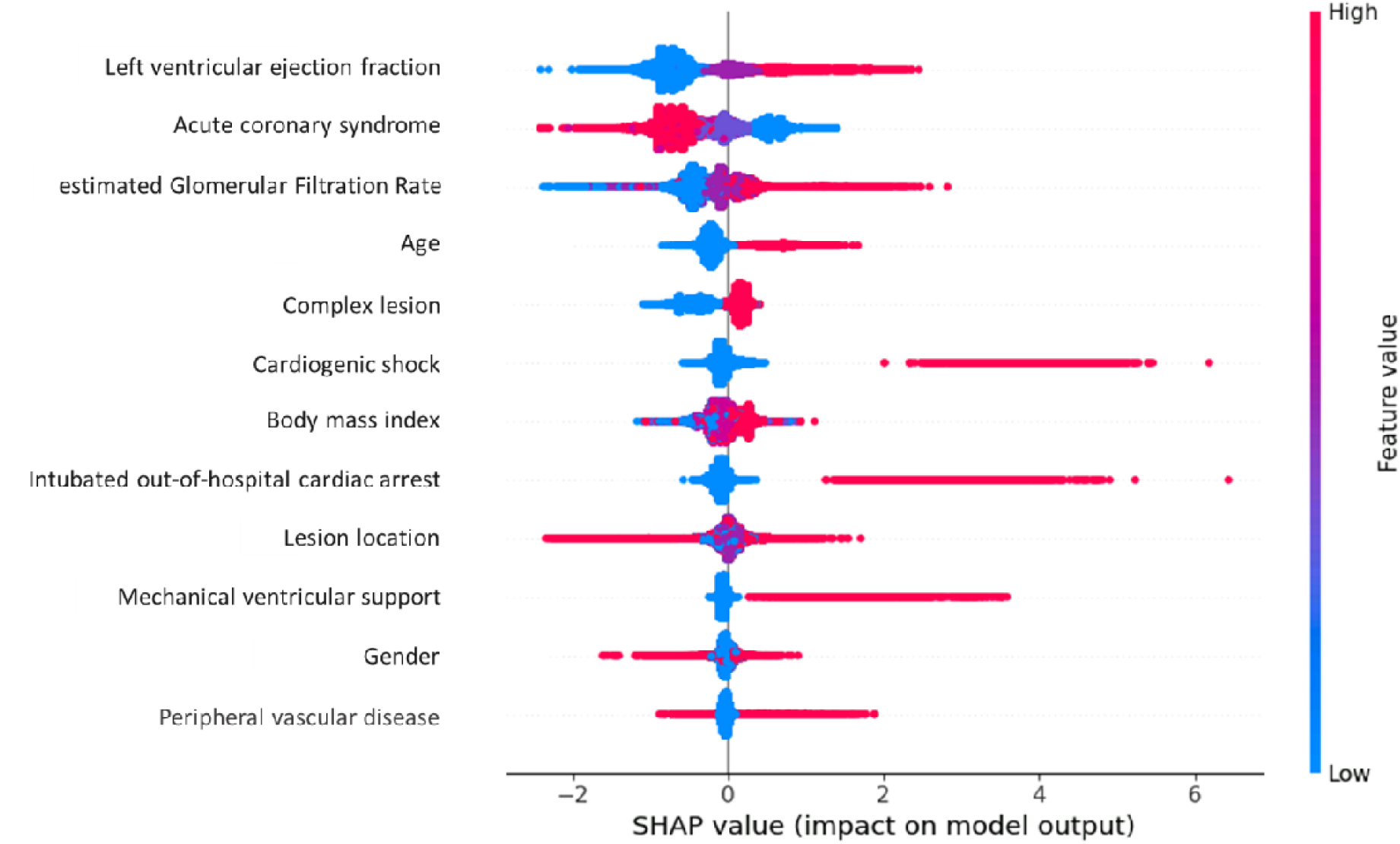
SHAP Beeswarm plot for XGB model to select top factors Note: SHAP, SHapley Additive exPlanations, XGB, Extreme Gradient Boosting Interpretation of the SHAP plot: The horizontal axis displays SHAP values, which represent the contribution of each variable to the model’s output. Negative SHAP values are associated with a lower likelihood of mortality, while positive values indicate a higher risk. The dots’ colours reflect the variable’s value for individual instances: blue indicates lower values, while red indicates higher values. The vertical bar on the right side provides the coding for categorical variables or the scale for numerical ones. For example, LVEF is coded as 0 for normal and 1 for mild, 2 for moderate and 3 for severely reduced ejection fraction (Table S1). Thus, blue dots represent normal values (code 0), while red dots correspond to severely reduced ejection fraction (code 3).

In Figure 4, the variables are ranked in order of importance, with those at the top having the greatest impact on the predictions. In the XGB model, LVEF was selected as the most influential predictor followed by ACS and eGFR. The plot also showed that severely reduced ejection fraction is the most influential predictors for 30-day all-cause mortality post-PCI. Other predictors of 30-day all-cause mortality included presentation with STEMI, severe renal impairment, age 80 years or older, B2/C lesion complexity, occurrence of cardiogenic shock, overweight or obesity, intubated OHCA, lesions located in the left main or graft vessels, use of mechanical ventricular support, female gender, and the presence of PVD (Figure 4).

### Health services’ performance comparison

The effectiveness of the XGB model in predicting 30-day all-cause mortality was evaluated by assessing the performance of health services using a funnel plot. In the funnel plot in Figure 5, one out of the 33 health services fell outside the 95.0% control limit when using the variables selected by the XGB model. However, none of the 33 health services exceeded the 99.8% control limit, indicating a consistent and reliable assessment of health service performance (Figure 5).

**Figure 5.**
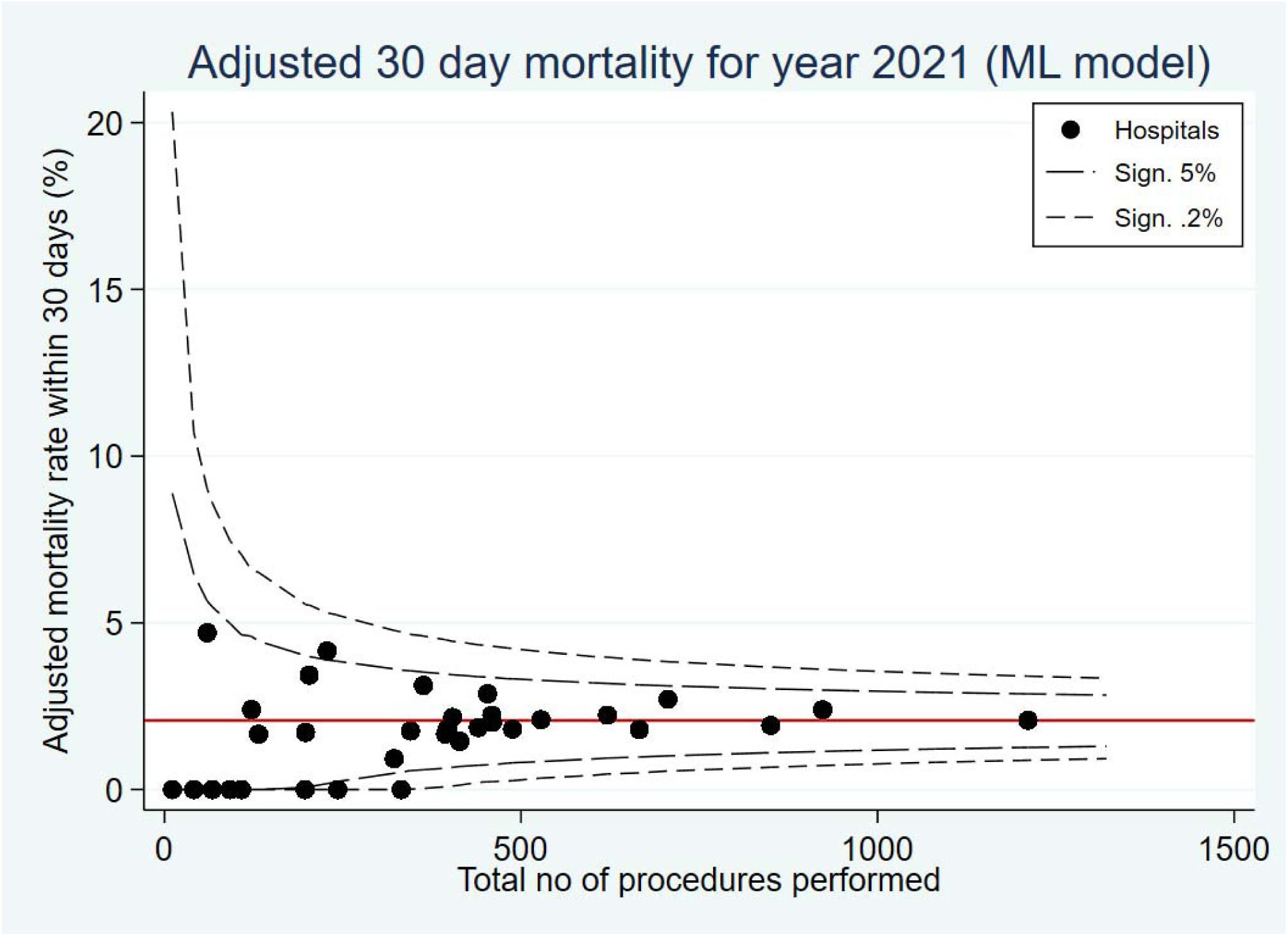
Funnel plot to assess health services’ performance Note: ML, Machine Learning; VCOR, Victorian Cardiac Outcome Registry.

## DISCUSSION

The primary aim of this study was to compare various ML models and identify an optimized risk-adjustment model for predicting 30-day all-cause mortality post-PCI. The XGB model demonstrated the best overall predictive performance and was selected as the optimal model. The model identified the key variables most strongly associated with mortality, allowing surgeons and patients to assess accurately the preoperative risk for individuals undergoing PCI.

Traditionally, the risk modelling predominately relies on traditional LR model.^7–10^ ^13^ ML models offer key advantages over LR models. Although ML does not always significantly outperform traditional models in accuracy, it excels in handling large, complex datasets by automatically detecting non-linearity and interaction without requiring manual input.^39^ In this study, seven most commonly used ML algorithms were employed to identify pre-procedural factors associated with 30-day all-cause mortality post-PCI. The XGB model, developed using 12 key factors selected by the Boruta method, and while applied to validation data set, it outperformed other models across most metrics, including accuracy, RMSE, specificity, precision, F1 score, and Brier score. Furthermore, it demonstrated excellent discrimination (ROC-AUC of 85.5%) and strong calibration, highlighting its robustness and reliability. However, a study from Taiwan found that the DT model demonstrated excellent predictive capability for 30-day mortality post-PCI,^36^ while in the United States, an Adaptive Booster classifier was identified as the optimal model for predicting various cardiac intervention outcomes.^35^ Furthermore, in an Italian population, the RF model was identified as superior in predicting 30-day all-cause mortality post-PCI.^40^ The variation in identified models across studies may be due to variation in population, differences in variable definition and availability of variable in the registries/database among others.^28^ Despite this variation, the findings suggest that ML-based models hold significant promise in developing predictive models for 30-day all-cause mortality post-PCI due to their high performance, capacity to handle large datasets, and ability to reduce the risk of overfitting in the presence of highly imbalanced outcome.^18–20^

The XGB model emphasized the pivotal role of LVEF as a primary factor significantly associated with 30-day all-cause mortality post-PCI. This aligns with previous research across diverse studies, underscoring LVEF as a critical factor linked to 30-day mortality post-PCI and affirming the robust performance of the model.^8^ ^10^ ^41^ In addition to LVEF, the XGB model identified ACS, eGFR, age, and complex lesion as the most influential factors associated with 30-day all-cause mortality post-PCI. These factors represent key patient-centered clinical and procedural characteristics that significantly influence post-PCI outcomes. Understanding them is clinically valuable for enabling early risk stratification, guiding informed clinical decisions, and supporting personalized care planning for patients undergoing PCI.^42–45^ A previous study conducted in Taiwan using a DT model identified hyperlipidemia, hypertension, diabetes, heart failure, stroke, and chronic kidney disease as factors associated with 30-day mortality post-PCI.^36^ It is worth noting that studies employing ML techniques to identify factors associated with 30-day mortality post-PCI remain limited within the Australian population context. However, an Australian study using a traditional LR approach (the VCOR risk adjustment model) identified nearly identical predictors to those found by the ML-based XGB model in the current study.^6^ Furthermore, consistent findings across multiple studies utilizing traditional LR models have underscored the importance of factors such as age, gender, BMI, LVEF, eGFR, cardiac arrest, ACS, mechanical ventricular support, PVD, CVD, and complex lesions as significant predictors of 30-day all-cause mortality post-PCI.^6^ ^10^ ^28^The current study did not include the percutaneous entry site, a variable featured in the VCOR model, which is specific to interventional cardiologists. However, this study focused on assessing patient-centred factors and deliberately excluded treatment-related variables to avoid compromising the accuracy of individual risk estimation in the risk adjustment model.

Risk-adjusted 30-day all-cause mortality prediction is disseminated to participating hospitals in the VCOR network, allowing for health service performance benchmarking against other health services via risk-adjusted funnel plots. The XGB model’s predictive ability in evaluating 30-day all-cause mortality post-PCI demonstrated encouraging results in assessing health service performance. The XGB model’s identification of critical risk factors for 30-day all-cause mortality post-PCI suggests the possibility of integrating within the VCOR risk-adjustment process and public reporting mechanisms. It may be beneficial to repeat these analyses when data over a longer period of time is available to reassess the presence of consistent differences in prediction.

### Strengths and limitations

The current study unveiled several strengths and limitations. The inclusion of a large volume of data substantially contributed in enhancing the accuracy of predicting 30-day all-cause mortality post-PCI is the key strength. Another strength of this study is the ML model’s ability to rank the most influential variables associated with mortality. The proposed set of pre-procedural variables holds promise for boosting the model’s overall performance. However, the study is not without its limitations. Firstly, the data source encompassed patients from a specific geographic region in Australia, constraining the generalizability of the findings and necessitating validation in diverse populations. Secondly, this study focused solely on patient-level risk and, therefore, included only patient-centred variables, excluding treatment choices and health system-related factors. The study only explored a subset of ML models, leaving unaddressed the performance of methods that were not evaluated herein. ML models are not able to produce p-value (significance level) and beta-coefficient while selecting influential variables. Finally, it’s worth noting that due to the limitations of the availability of all 17 factors, such as hypertension, single or multivessel diseases, heart failure, Thrombolysis in Myocardial Infarction (TIMI) flow and urgency of PCI, proposed as per earlier study,^28^ that there may be missed opportunities for improving overall performance metrics.

## CONCLUSION

In this study, seven ML based risk prediction models for 30-day all-cause mortality post-PCI were developed and compared. The XGB model was selected as the best performing model. This model selected LVEF, ACS, eGFR, age, and complex lesion as the top influential factors. This model has the potential to assist clinicians in early identification of patients with high risk factors and benchmarking health service performance. However, there is need for further validation utilising external data to ensure the applicability of these findings to other patients’ population.

## Data Availability

All data produced in the present study are available upon reasonable request to the authors

## Abbreviations

ACS: Acute coronary syndrome
AUC: Area under the curve
BMI: Body mass index
CABG: coronary artery bypass grafting
CI: Confidence interval
CTO: Chronic total occlusion
CVD: Cerebrovascular disease
DT: Decision Tree
eGFR: estimated glomerular filtration rate
GB: Gradient Booster
LDA: Linear Discriminatory Analysis
LR: Logistic Regression
MICE: Multiple Imputations by Chained Equations
ML: Machine Learning
OHCA: Out-of-hospital cardiac arrest
PCI: Percutaneous coronary intervention
PVD: Peripheral vascular disease
RF: Random Forest
ROC: Receiver operating characteristic
SGB: Stochastic Gradient Boosting
SHAP: SHapley Additive exPlanations
STEMI: ST-elevated myocardial infraction
TIMI: Thrombolysis in Myocardial Infarction
VCOR: Victorian cardiac outcome registry
XGB: Extreme Gradient Booster

## Contributors

Conceptualisation: MRKC, BB, DD, DS. Methodology: BB, MRKC, MNK. Analysis: MRKC, MAM, MSI. Manuscript drafting: MRKC. Manuscript review and critical revision: BB, DS, DD, AB, CMR, SN, JL, DPC. Visualisation: MRKC, MSI. Supervision: BB, DS, DD, MNK. Project administration: DS, DD. MRKC, BB are the guarantor.

## Patient consent for publication

Not applicable.

## Ethics approval

The VCOR was primarily approved by the ethics committee at The Alfred Hospital, Melbourne, Australia (approval number 47/12), and also approved by each participating hospital, including the use of opt-out consent. The current project received ethical approval from the Monash University Human Research Ethics Committee (MUHREC), under the project reference number 2022-35388-80724.

## Data availability statement

Data are available upon reasonable request. Anonymized personal data were obtained from the Victorian Cardiac Outcome Registry (VCOR) after ethical approval and a confidentiality assessment. In accordance with Australian laws and regulations, access to personal sensitive data is restricted to researchers who meet the legal requirements for such access. For inquiries regarding data access, please contact Dr. Diem Dinh.

## References

1. Khera S, Kolte D, Bhatt DL. Percutaneous coronary intervention. Translational research in coronary artery disease: Elsevier 2016:179–94.

2. Jennings S, Bennett K, Shelley E, et al. Trends in percutaneous coronary intervention and angiography in Ireland, 2004–2011: implications for Ireland and Europe. IJC Heart & Vessels 2014;4:35–39.

3. Virani SS, Alonso A, Benjamin EJ, et al. Heart disease and stroke statistics—2020 update: a report from the American Heart Association. Circulation 2020;141(9):e139–e596.

4. AIHW. Heart, stroke and vascular disease: Australian facts. National Hospital Morbidity Database (NHMD). Australian Institute of Health and Welfare (AIHW). 2020

5. Serruys PW, Morice M-C, Kappetein AP, et al. Percutaneous coronary intervention versus coronary-artery bypass grafting for severe coronary artery disease. New England journal of medicine 2009;360(10):961–72.

6. Tacey M, Dinh DT, Andrianopoulos N, et al. Risk-adjusting key outcome measures in a clinical quality PCI registry: development of a highly predictive model without the need to exclude high-risk conditions. JACC: Cardiovascular Interventions 2019;12(19):1966–75.

7. Andrews M, Iqbal J, Wall JJ, et al. Development and Validation of a Novel Risk Score for Primary Percutaneous Coronary Intervention for ST-Elevation Myocardial Infarction. Cardiovascular Revascularization Medicine 2019;20(11):980–84.

8. Bulluck H, Zheng H, Chan MY, et al. Independent predictors of cardiac mortality and hospitalization for heart failure in a multi-ethnic Asian ST-segment elevation myocardial infarction population treated by primary percutaneous coronary intervention. Scientific Reports 2019;9(1):10072.

9. Cheng JM, Helming AM, van Vark LC, et al. A simple risk chart for initial risk assessment of 30-day mortality in patients with cardiogenic shock from ST-elevation myocardial infarction. European Heart Journal: Acute Cardiovascular Care 2016;5(2):101–07.

10. Cockburn J, Kemp T, Ludman P, et al. Percutaneous coronary intervention in octogenarians: a risk scoring system to predict 30-day outcomes in the elderly. Catheterization and Cardiovascular Interventions 2021;98(7):1300–07.

11. Goldstein BA, Navar AM, Pencina MJ, et al. Opportunities and challenges in developing risk prediction models with electronic health records data: a systematic review. Journal of the American Medical Informatics Association: JAMIA 2017;24(1):198.

12. McCreanor V, Nowbar A, Rajkumar C, et al. Cost-effectiveness analysis of percutaneous coronary intervention for single-vessel coronary artery disease: an economic evaluation of the ORBITA trial. BMJ open 2021;11(2):e044054.

13. Marcolino MS, Simsek C, De Boer SP, et al. Short-and long-term major adverse cardiac events in patients undergoing percutaneous coronary intervention with stenting for acute myocardial infarction complicated by cardiogenic shock. Cardiology 2012;121(1):47–55.

14. Panch T, Pearson-Stuttard J, Greaves F, et al. Artificial intelligence: opportunities and risks for public health. The Lancet Digital Health 2019;1(1):e13–e14.

15. Aldridge RW. Research and training recommendations for public health data science. The Lancet Public Health 2019;4(8):e373.

16. Motwani M, Dey D, Berman DS, et al. Machine learning for prediction of all-cause mortality in patients with suspected coronary artery disease: a 5-year multicentre prospective registry analysis. European heart journal 2017;38(7):500–07.

17. Breiman L. Statistical modeling: The two cultures (with comments and a rejoinder by the author). Statistical science 2001;16(3):199–231.

18. Goldstein BA, Navar AM, Pencina MJ, et al. Opportunities and challenges in developing risk prediction models with electronic health records data: a systematic review. Journal of the American Medical Informatics Association 2017;24(1):198–208.

19. Shickel B, Tighe PJ, Bihorac A, et al. Deep EHR: a survey of recent advances in deep learning techniques for electronic health record (EHR) analysis. IEEE journal of biomedical and health informatics 2017;22(5):1589–604.

20. Risk prediction with electronic health records: A deep learning approach. Proceedings of the 2016 SIAM international conference on data mining; 2016. SIAM.

21. Morgan DJ, Bame B, Zimand P, et al. Assessment of machine learning vs standard prediction rules for predicting hospital readmissions. JAMA network open 2019;2(3):e190348–e48.

22. Kehl KL, Elmarakeby H, Nishino M, et al. Assessment of deep natural language processing in ascertaining oncologic outcomes from radiology reports. JAMA oncology 2019;5(10):1421–29.

23. Dagliati A, Marini S, Sacchi L, et al. Machine learning methods to predict diabetes complications. Journal of diabetes science and technology 2018;12(2):295–302.

24. Peng Y, Du X, Rogers KD, et al. Predicting in-hospital mortality in patients with acute coronary syndrome in China. The American Journal of Cardiology 2017;120(7):1077–83.

25. Shouval R, Hadanny A, Shlomo N, et al. Machine learning for prediction of 30-day mortality after ST elevation myocardial infraction: An Acute Coronary Syndrome Israeli Survey data mining study. International journal of cardiology 2017;246:7–13.

26. Kwon J-m, Jeon K-H, Kim HM, et al. Deep-learning-based out-of-hospital cardiac arrest prognostic system to predict clinical outcomes. Resuscitation 2019;139:84–91.

27. Lefkovits J BA, Dinh D, Carruthers H, Doyle J, Lucas M, Stub D, Reid CM The Victorian Cardiac Outcomes Registry Annual Report 2021 Monash University, SPHPM August 2022, Report No 9, pages 8. 2021

28. Chowdhury MRK, Stub D, Dinh D, et al. Preoperative Variables of 30-Day Mortality in Adults Undergoing Percutaneous Coronary Intervention: A Systematic Review. Heart, Lung and Circulation 2024

29. Xu B-Z, Wang B, Chen J-P, et al. Construction and validation of a personalized risk prediction model for in-hospital mortality in patients with acute myocardial infarction undergoing percutaneous coronary intervention. Clinics 2025;80:100580.

30. Liu Y, De A. Multiple imputation by fully conditional specification for dealing with missing data in a large epidemiologic study. International journal of statistics in medical research 2015;4(3):287.

31. Hair JF, Black WC, Babin BJ, et al. Multivariate data analysis: Cengage Learning EMEA, 2019.

32. Chen R-C, Dewi C, Huang S-W, et al. Selecting critical features for data classification based on machine learning methods. Journal of Big Data 2020;7(1):52.

33. Lundberg SM, Lee S-I. A unified approach to interpreting model predictions. Advances in neural information processing systems 2017;30

34. Shetty MK, Kunal S, Girish M, et al. Machine learning based model for risk prediction after ST-Elevation myocardial infarction: Insights from the North India ST elevation myocardial infarction (NORIN-STEMI) registry. International journal of cardiology 2022;362:6–13.

35. Al’Aref SJ, Singh G, van Rosendael AR, et al. Determinants of In-Hospital Mortality After Percutaneous Coronary Intervention: A Machine Learning Approach. J Am Heart Assoc 2019;8(5):e011160. doi: 10.1161/jaha.118.011160 [published Online First: 2019/03/06]

36. Hsieh MH, Lin SY, Lin CL, et al. A fitting machine learning prediction model for short-term mortality following percutaneous catheterization intervention: a nationwide population-based study. Ann Transl Med 2019;7(23):732. doi: 10.21037/atm.2019.12.21 [published Online First: 2020/02/12]

37. Spiegelhalter DJ. Funnel plots for comparing institutional performance. Statistics in medicine 2005;24(8):1185–202.

38. Yan R, Zhang H, Shi B, et al. Sex Disparities in In-Hospital Outcomes After Percutaneous Coronary Intervention (PCI) in Patients With Acute Myocardial Infarction and a History of Coronary Artery Bypass Grafting (CABG): A Cross-Sectional Study. Health Science Reports 2024;7(12):e70292.

39. Christodoulou E, Ma J, Collins GS, et al. A systematic review shows no performance benefit of machine learning over logistic regression for clinical prediction models. J Clin Epidemiol 2019;110:12–22. doi: 10.1016/j.jclinepi.2019.02.004 [published Online First: 2019/02/15]

40. Burrello J, Gallone G, Burrello A, et al. Prediction of all-cause mortality following percutaneous coronary intervention in bifurcation lesions using machine learning algorithms. Journal of Personalized Medicine 2022;12(6):990.

41. Hamburger JN, Walsh SJ, Khurana R, et al. Percutaneous coronary intervention and 30-day mortality: The British Columbia PCI risk score. Catheterization and Cardiovascular Interventions 2009;74(3):377–85.

42. Shahian DM, O’Brien SM, Filardo G, et al. The Society of Thoracic Surgeons 2008 cardiac surgery risk models: part 1--coronary artery bypass grafting surgery. Ann Thorac Surg 2009;88(1 Suppl):S2–22. doi: 10.1016/j.athoracsur.2009.05.053 [published Online First: 2009/07/09]

43. O’Gara PT, Kushner FG, Ascheim DD, et al. 2013 ACCF/AHA guideline for the management of ST-elevation myocardial infarction: a report of the American College of Cardiology Foundation/American Heart Association Task Force on Practice Guidelines. J Am Coll Cardiol 2013;61(4):e78-e140. doi: 10.1016/j.jacc.2012.11.019 [published Online First: 2012/12/22]

44. Endo A, Kawamura A, Miyata H, et al. Angiographic Lesion Complexity Score and In-Hospital Outcomes after Percutaneous Coronary Intervention. PLoS One 2015;10(6):e0127217. doi: 10.1371/journal.pone.0127217 [published Online First: 2015/06/30]

45. Peerwani G, Khan SM, Khan MD, et al. Gender Differences in Clinical Outcomes After Percutaneous Coronary Intervention-Analysis of 15,106 Patients from the Cardiac Registry of Pakistan Database. Am J Cardiol 2023;188:61–67. doi: 10.1016/j.amjcard.2022.11.020 [published Online First: 2022/12/07]

