## Supplemental Table 1-5, Figure 1-3 for "Risk Prediction Modelling of 30-day all-cause mortality following percutaneous coronary intervention in an Australian population: Leveraging Machine Learning"

**Table S1** Operational definition of preoperative exposure variables

| **Variables** | **Definition** | **Scale of measurement and code** | **Type** |
| --- | --- | --- | --- |
| Age (group in years) | The date of birth of the patients was collected in DD/MM/YYYY format. | <80 years (0), ≥80 years (1) | Binary |
| Gender | The gender (sex) of the patients | Male (0), female (1) | Binary |
| Body mass index (BMI) | Body mass index was calculated using following formula, BMI=Weight in kg / (Height in meter)^2^ kg/m^2^.  Weight was measured in Kg in light clothing and height in centimetre in bare or stockinged feet. Height and weight measurement could be self-reported. | Underweight (<18.5 kg/m2) (1), normal (18.5-24.9 kg/m2) (0), Overweight (25.0-29.9 kg/m2) (2), obesity (30.0 kg/m2 and above) (3) | Categorical |
| Acute coronary syndrome (ACS) | ACS encompasses clinical features comprising chest pain or overwhelming shortness of breath, defined by accompanying clinical, ECG and biochemical features. Specifically, ACS refers to unstable angina, non-ST-Elevation Myocardial Infarction (NSTEMI) and/or ST-Elevation Myocardial Infarction (STEMI). The patient must have experienced an ACS event within the last 7 days to be  coded “yes” for ACS at the time of the PCI event. | STEMI (1), NSTEMI (2), unstable angina (3), no-ACS (4) | Categorical |
| Cardiogenic shock | Cardiogenic shock is coded as ‘yes’ if all of the following apply:  1. Sustained (>30 minutes) episode of systolic blood pressure <90 mm Hg (or  vasopressors required to maintain BP >90 mm Hg); AND  2. Evidence of elevated filling pressures (e.g. pulmonary congestion on examination or chest radiograph); AND  3. Evidence of end organ hypoperfusion (e.g. urine output 30mL/hour; or  cold/diaphoretic extremities; or altered mental status, etc.). | No (0), yes (1) | Binary |
| Intubated out-of-hospital cardiac arrest (IOHCA) | Cardiac arrest is coded as ‘yes’ if any one of the following apply:  1. if the patient has experienced an out of hospital cardiac arrest (i.e. the lack of effective cardiac output), including if the person was under cardiac arrest at the time of presentation to the hospital and/or the patient was intubated prior to the PCI procedure. and  2. if the patient was intubated prior to the PCI procedure. | No (0), yes (1) | Binary |
| Left Ventricular Ejection Fraction (LVEF) | For all patients (excluding STEMIs) indicate whether the patient’s  ventricular ejection fraction (EF) was measured (or estimated) within 6 months prior to the current procedure and up to four weeks post-discharge.  This includes the period leading up to and including the cardiac catheter lab visit, after the lab visit and up to four weeks after the patient was discharged. If multiple test results are available during this period, select the test result closest to the date/time of the current PCI.  For STEMI patients a LVEF test must have been recorded during the current  admission or up to four weeks post discharge for this item to be coded ‘yes’.  For these patients, where no LVEF test was performed during the current admission or up to four weeks post discharge, code “no”.  EF tests include, but are not exclusive to, angiography, echocardiography, nuclear  stress tests and imaging scans. | Normal (0), mild (1), moderate (2), severe (3) | Binary |
| estimated Glomerular Filtration Rate (eGFR) | Record the last serum creatinine levels recorded within 60 days prior to the current PCI  (in μmol/L). To convert from mmol/L to μmol/L multiply by 1000 or move decimal point 3 spaces to the right. The formula for eGFR uses age, gender and the level of creatinine in blood to estimate GFR.   - eGFR of 90 or higher is in the normal range. - eGFR of 60 -89 may mean early-stage kidney disease/mild. - eGFR of 30 -59 may mean kidney disease/moderate.   eGFR below 30 may mean kidney failure/severe | Normal (0), mild (1), moderate (2), severe (3) | Categorical |
| Mechanical ventricular support | If a patient required mechanical ventricular support prior to the current PCI procedure. | No (0), yes (1) | Binary |
| Diabetes | If the patient had a current medical diagnosis of diabetes that required medical intervention (regardless of duration of the disease). This includes a medical diagnosis made during the current admission where medication is prescribed.  The patient must be on anti-diabetic medication to lower blood sugar. Anti-  diabetic medications to lower blood sugar include oral hypoglycaemics and insulin. For patients whose diabetes is controlled with diet alone, code as ‘no’. | No (0), yes (1) | Binary |
| Peripheral vascular disease (PVD) | If the patient displays evidence of either chronic or acute PVD. The presence of  PVD must be demonstrated by vascular reconstruction or amputation for arterial  insufficiency, bypass surgery or percutaneous intervention. PVD includes the aorta, extremities and carotid vessels. | No (0), yes (1) | Binary |
| Cerebrovascular disease (CVD) | Indicate whether the patient has a history of stroke or cerebrovascular accident,  resulting from an ischaemic or intracerebral haemorrhagic event only where the patient suffered a loss of neurological function with residual symptoms remaining for at  least 72 hours. | No (0), yes (1) | Binary |
| Chronic total occlusion | Indicate whether the current lesion was presumed to be a chronic total occlusion. Chronic total occlusion  Is defined as being>3 months old and/or bridging collaterals. | No (0), yes (1) | Binary |
| Previous percutaneous coronary intervention (PCI) | If the patient has had a prior Percutaneous Transluminal Coronary Angioplasty,  Coronary Atherectomy, and/or coronary stent done at any time prior to the current PCI procedure. | No (0), yes (1) | Binary |
| Previous coronary artery bypass grafting (CABG) | If the patient has had a prior CABG. | No (0), yes (1) | Binary |
| Lesion complexity | Lesion type according to the ACC/AHA classification guideline for current lesion.  Type A: Minimally complex, discrete (<10 mm), concentric, readily accessible, lesion in non-angulated segment (<45 degrees), smooth contour, little or no calcification, less than totally occlusion, not ostial in location, no major side branch involvement, absence of thrombus.  Type B: Only one type B characteristics - lesion moderately complex, tubular (10-20 mm), eccentric, moderately tortuosity of proximal segments, lesion in moderately angulated segment (>45 degrees but <90 degrees), irregular contour, moderately to heavy calcification, total occlusion less than three months old, ostial in location, bifurcation lesions requiring double guide wires, some thrombus present.  Type B2: More than one type B characteristics  Type C: severe complex diffuse (>20mm), excessive tortuosity of proximal segment, lesion in extremely angulated segment > 90 degrees, total occlusion greater than 3 months old or bridging collaterals, inability to protect major side branches, degenerated vein graft with friable lesion. | Type A and B (0),  and Type B2 and C (1) | Binary |
| Lesion location | Indicate the coronary segment that applies for each coronary lesion attempted during the current PCI. Every coronary lesion attempted during the current PCI must be recorded separately. Up to five lesions per PCI can be recorded.   - Right coronary artery - Left anterior descending - Circumflex artery - Left main - Graft | Right coronary artery (1), left anterior descending (2), circumflex artery (3), left main (4), graft (5) | Categorical |

**Table S2** Short description of selected machine learning methods

| **Items** | **Description** |
| --- | --- |
| Decision Tree (DT) | A DT machine learning model is a predictive model that uses a tree-like structure of decisions and their possible outcomes to make predictions or classifications. The tree is constructed based on features of the data, and each internal node represents a decision based on a specific feature, leading to branches representing possible outcomes. The leaves of the tree represent the final predicted class or outcome.  DTs are popular for their interpretability and ease of visualization. The model is built through a process of recursively splitting the data into subsets based on the most informative features. The decision-making process follows a path from the root to the leaves, where each split is determined by optimizing criteria such as Gini impurity or information gain. |
| Gradian Booster (GB) | A GB machine learning model is an ensemble technique that builds a strong predictive model by combining the outputs of multiple weak learners, typically decision trees. Unlike traditional boosting methods, gradient boosting optimizes the model's performance by minimizing the errors of the previous weak learners.  Gradient boosting works in an iterative manner, where each weak learner is trained to correct the errors of the ensemble's current predictions. The model assigns more weight to instances that were previously misclassified or had higher errors. Subsequent weak learners focus on these challenging instances, gradually improving the overall model's accuracy.  This model is widely used in both regression and classification tasks and are known for their ability to handle complex relationships in data, robustness, and high predictive performance |
| Linear Discriminant Analysis (LDA) | LDA is a supervised machine learning method used for classification and dimensionality reduction. It finds a linear combination of features that maximizes class separability by maximizing between-class variance and minimizing within-class variance. LDA assumes normal distribution of classes and works best with linearly separable data. It is widely applied in face recognition, medical diagnosis, and finance. |
| Logistic Regression (LR) | LR is a supervised machine learning model used for binary and multiclass classification. It predicts probabilities using the sigmoid function and optimizes weights through maximum likelihood estimation. The model assumes a linear relationship between features and the log-odds of the target. Regularization techniques (L1/L2) help prevent overfitting. |
| Random Forest (RF) | The RF model is an ensemble algorithm that constructs multiple decision trees during training. Each tree is built using a random subset of features and data samples, and the final prediction is determined by aggregating the outputs of these individual trees. Known for its robustness and ability to handle complex datasets, Random Forest is widely used in classification and regression tasks, offering improved accuracy and resistance to overfitting compared to individual decision trees. Additionally, it provides insights into feature importance, making it a versatile and effective tool in various applications. |
| Stochastic Gradient Boosting (SGB) | SGB is an ensemble learning method that enhances Gradient Boosting by incorporating randomness to improve generalization and reduce overfitting. It builds decision trees sequentially, with each tree correcting the errors of the previous ones. Unlike standard Gradient Boosting, SGB randomly samples a subset of the training data (stochastic sampling) before fitting each tree, which introduces diversity and improves model robustness. This method is widely used for structured data, due to its strong predictive performance. |
| Extreme Gradient Booster (XGB) | The XGB model is a powerful machine learning algorithm designed for both classification and regression tasks. It belongs to the ensemble learning category and is an extension of the gradient boosting framework. XGB excels in performance due to its efficient implementation and optimization techniques. It sequentially builds a series of decision trees, each correcting the errors of the previous one, resulting in a robust and accurate model. Notable features include regularization to prevent overfitting, handling missing values, and the ability to process large datasets quickly. XGB has gained popularity for its versatility, speed, and outstanding predictive performance in various real-world applications. |

**Table S3.** Model optimization

| **Items** | **Hyperparameter optimization** | **Cross-validation (k-fold)** | **Parameters** | **Thresholds** |
| --- | --- | --- | --- | --- |
| Decision Tree (DT) | Grid search | 10 folds | criterion, max_depth, min_samples_split,  min_sample_leaf, splitter, random_state | 0.4 |
| Gradian Booster (GB) | Grid search | 10 folds | n_estimators, min_samples_split,  min_sample_leaf,  learning_rate, learning_rate, max_features, sub_smaple, scoring, n_jobs, random_state | 0.4 |
| Linear Discriminant Analysis (LDA) | Grid search | 10 folds | solver, random_state | 0.4 |
| Logistic Regression (LR) | Grid search | 10 folds | random_state, C, penalty, solver, scoring | 0.4 |
| Random Forest (RF) | Grid search | 10 folds | n_estimators, min_samples_split,  max_features | 0.4 |
| Stochastic Gradient Boosting (SGB) | Grid search | 10 folds | log_loss, random_state, penalty, alpha, l1_ratio | 0.4 |
| Extreme Gradient Booster (XGB) | Grid search | 10 folds | random_state, objective, alpha, gamme, eval_metrics, eta, learning_rate, max_delta_step, nthread, n_estimators, max_depth, min_child_weight, colsample_bytree, colsample_bylevel, scoring | 0.4 |

**Table S4.** Defining performance metrics

| True positive | A true positive is an outcome where the model correctly predicts the positive class |
| --- | --- |
| True negative | A true negative is an outcome where the model correctly predicts the negative class |
| False positive | A false positive is an outcome where the model incorrectly predicts the positive class |
| False negative | a false negative is an outcome where the model incorrectly predicts the negative class. |
| Accuracy | Model accuracy is defined as the number of classifications a model correctly predicts divided by the total number of predictions made which can be represented as: Accuracy = (True positives + True negatives) / (True positives + True negatives + False positives + False negatives). |
| Root Mean Square Error (RMSE) | RMSE=$\sqrt{\frac{1}{n}\sum_{i=1}^{n} {{(y}_{i}-\hat{y}_{i})}^{2}}$  RMSE quantifies the average prediction error, with greater emphasis on larger errors. A lower RMSE indicates better model accuracy |
| Sensitivity/recall | Sensitivity refers to a test's ability to designate an individual with disease as positive. A highly sensitive test means that there are few false negative results, and thus fewer cases of disease are missed. Sensitivity = true positive /(false negative + true positive) |
| Specificity | Specificity measures the proportion of true negatives that are correctly identified by the model. Specificity = true negative / (true negative + false positive) |
| Precision | The formula for precision (also called positive predictive value, PPV) is:  Precision = True positive/(True positive + False positive)  It measures the proportion of correctly predicted positive cases out of all predicted positive cases |
| F1 score | The **F1 score** is the harmonic mean of **precision** and **recall,** balancing both metrics. It is calculated as:  F1=2*(Precision*recall)/(Precision + Recall)  The F1 score provides a balance between precision and recall, making it useful when false positives and false negatives have similar consequences.  Interpretation:   - F1 = 1 → Perfect precision and recall (ideal model). - F1 = 0 → The model has either zero precision or zero recall (worst performance). - Higher F1 Score → Better model performance, meaning it effectively identifies positive cases while minimizing false positives and false negatives. - Lower F1 Score → Poor performance, indicating the model struggles with either false positives or false negatives.   The F1 score is crucial for imbalanced datasets, where accuracy alone can be misleading. |
| Receiver Operating Characteristics (ROC) | The plot of sensitivity versus 1-Specifity is called receiver operating characteristic (ROC) curve and the area under the curve (AUC). The mathematical formula of AUC is as follows  ROC*=*$\int_{x=0}^{1} [Sensitivity \{\left( 1-Specificity \right)^{-1} \left( x \right)\}]dx$ |
| Precision-Recall (PR) curve | The PR Curve is used to evaluate the performance of classification models, particularly in imbalanced datasets. It plots Precision (the accuracy of positive predictions) against Recall (the ability to identify all positives) at various thresholds. A high Precision and Recall indicate a strong model, while a low Precision and Recall suggest poor performance. The AUC-PR score summarizes model effectiveness, with a higher value indicating better performance. |
| Brier score | The **Brier Score** is a measure of the accuracy of probabilistic predictions. It evaluates how close the predicted probabilities are to the actual binary outcomes.  Brier score = 1/N$\sum_{i=1}^{N} {(f_{i}-o_{i})}^{2}$  Where:   - N = Total number of predictions - f_i_ = Predicted probability for instance iii - o_i_ = Actual outcome (1 for positive, 0 for negative)   The Brier Score ranges from 0 to 1, where **0** indicates perfect predictions, **1** represents completely incorrect predictions, and lower scores signify better calibration and accuracy of probabilistic predictions. |
| Calibration plot | A calibration plot visualizes the alignment between predicted probabilities and actual outcomes, with the x-axis representing predicted probabilities and the y-axis showing observed event proportions. A diagonal line indicates perfect calibration, while deviations from this line reveal miscalibration: predictions above the line suggest underestimation, and those below the line indicate overestimation. |

**Table S5.** Models’ performance (train/development dataset) in predicting 30-day mortality post-PCI

| **Models** | **Performance metrices of models** | | | | | | |
| --- | --- | --- | --- | --- | --- | --- | --- |
|  | **Accuracy** | **RMSE** | **Sensitivity/recall** | **Specificity** | **Precision** | **F1 score** | **Brier score** |
| **DT** | 0.872 | 0.358 | 0.917 | 0.821 | 0.537 | 0.520 | 0.128 |
| **GB** | 0.827 | 0.415 | 0.872 | 0.764 | 0.535 | 0.450 | 0.173 |
| **LDA** | 0.797 | 0.451 | 0.808 | 0.768 | 0.536 | 0.503 | 0.203 |
| **LR** | 0.798 | 0.449 | 0.826 | 0.749 | 0.535 | 0.494 | 0.202 |
| **RF** | 0.853 | 0.384 | 0.875 | 0.805 | 0.536 | 0.514 | 0.247 |
| **SGB** | 0.798 | 0.450 | 0.829 | 0.735 | 0.533 | 0.486 | 0.202 |
| **XGB** | 0.872 | 0.358 | 0.904 | 0.830 | 0.540 | 0.527 | 0.128 |

Note: RMSE, root mean square error; DT, Decision Tree; XGB, Extreme Gradient Boosting; GB, Gradient Boosting; LDA, Linear Discriminant Analysis; LR, Logistic Regression; RF, Random Forest; SGB, Stochastic Gradient Boosting

For Accuracy, Sensitivity, Specificity, Precision and F1 score, higher values indicate better model performance. For RMSE and Brier score lower values indicate better model performance

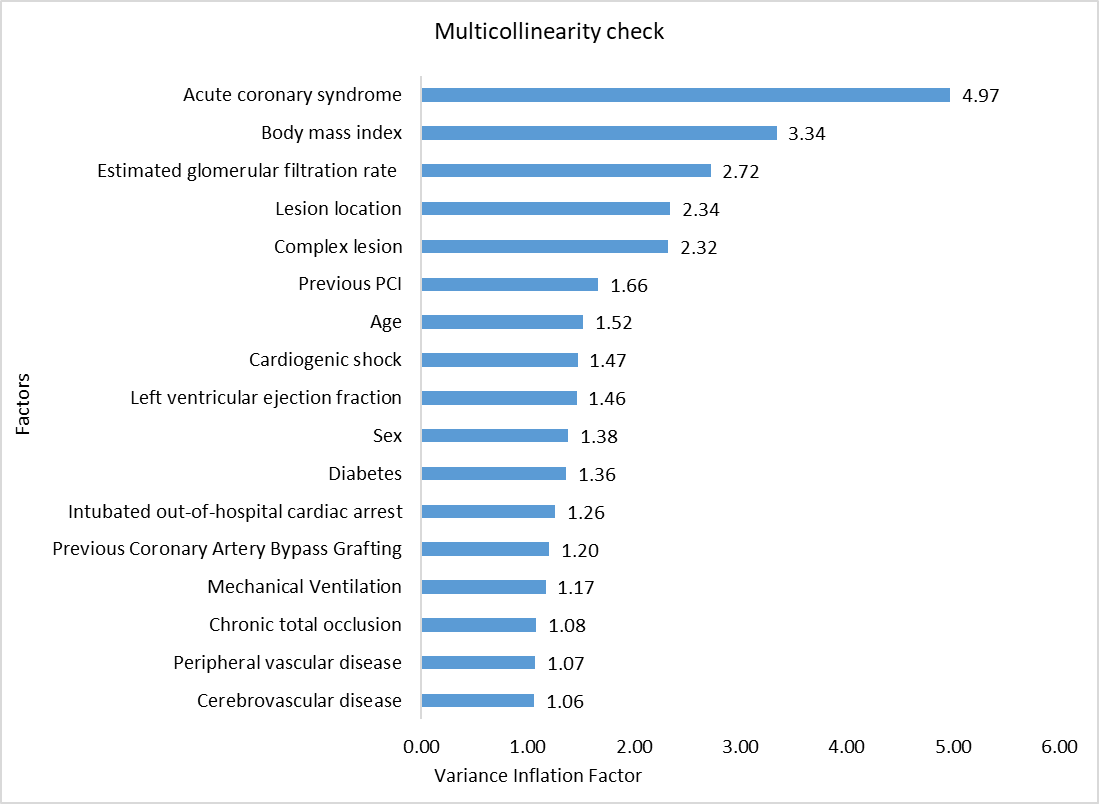

**Figure S1.** Multicollinearity checks of selected factors

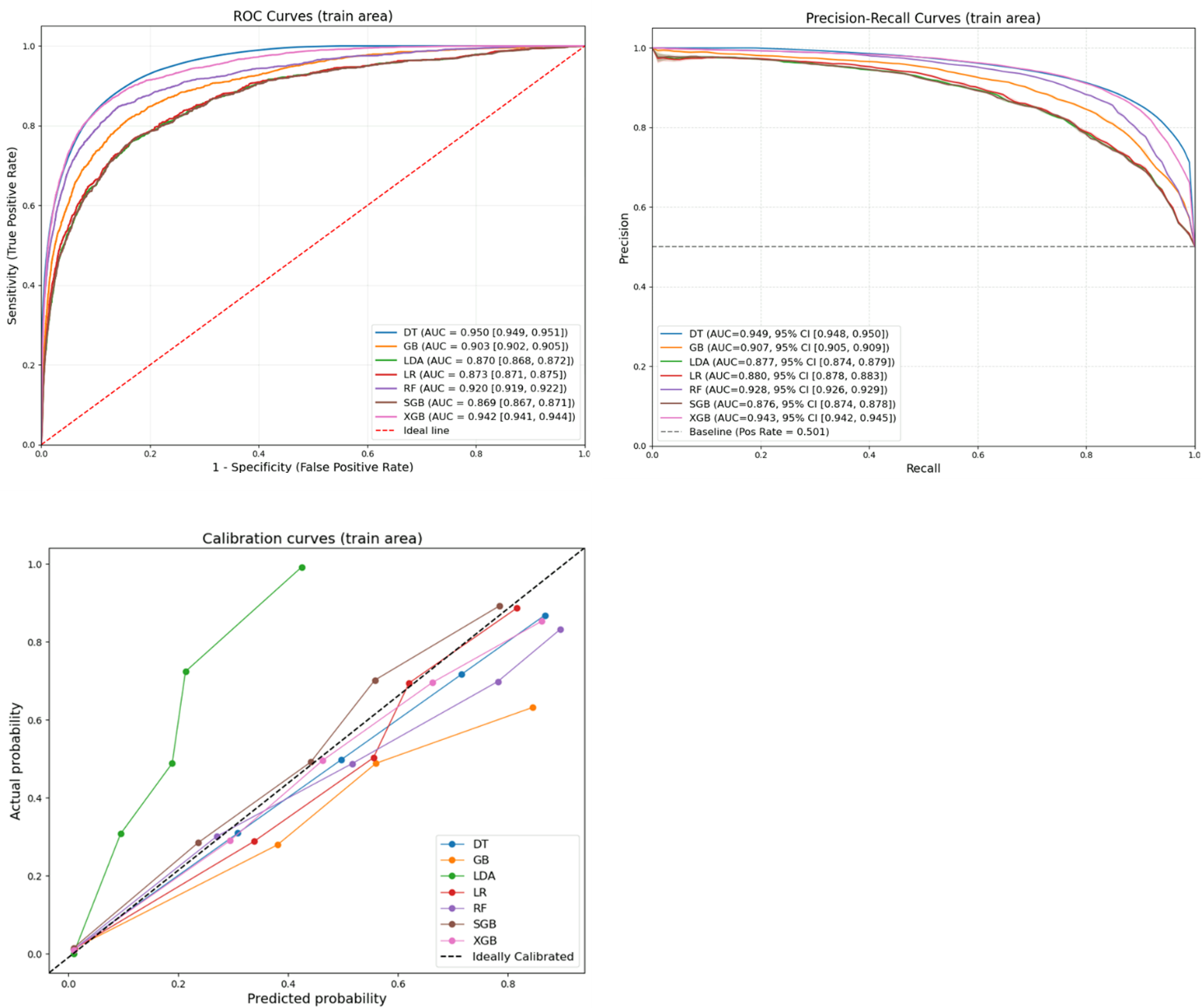

**Figure S2.** Receiver operating characteristics-area under the curve (ROC-AUC) curve, precision-recall curve and calibration curve for ML models (training/development dataset)

Note: CI, confidence interval; DT, Decision Tree; GB, Gradient Boosting; LDA, Linear Discriminant Analysis; LR, Logistic Regression; RF, Random Forest; SGB, Stochastic Gradient Boosting; XGB, Extreme Gradient Boosting.

For both ROC and PR scores, higher values indicate better model performance

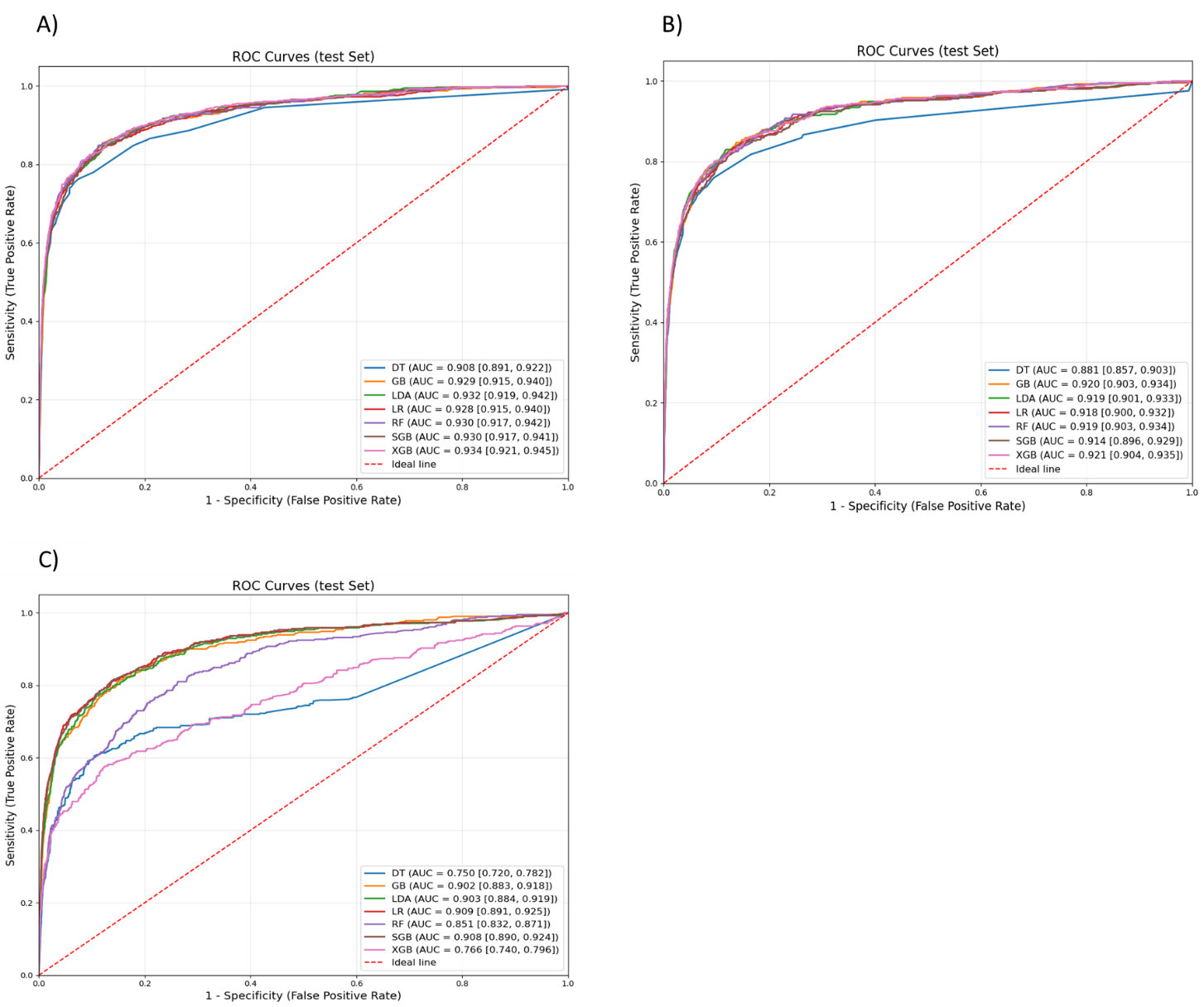

**Figure S3.** ROC-AUC curve for A) without addressing class imbalance B) dataset with missing values removed and without addressing class imbalance C) dataset with missing values removed and addressing class imbalance

Note: SHAP, SHapley Additive exPlanations, LR, Logistic Regression; SGB, Stochastic Gradient Boosting
